# Complementary non-sputum diagnostic testing using oral swabs and urine LAM testing for TB in people with HIV

**DOI:** 10.1101/2022.02.16.22271079

**Authors:** Adrienne E. Shapiro, Alaina M. Olson, Lara Kidoguchi, Xin Niu, Zinhle Ngcobo, Zanele P. Magcaba, Mduduzi W. Ngwane, Grant R. Whitman, Kris M. Weigel, Rachel C. Wood, Doug P.K. Wilson, Paul K. Drain, Gerard A. Cangelosi

**Author notes:** Corresponding author: Gerard Cangelosi, Dept. of Environmental and Occupational Health Sciences, Box 351618, University of Washington, Seattle, WA 98195.

## Abstract

Testing for mycobacterial lipoarabinomannan (LAM) in urine is a practical but insensitive alternative to sputum testing to diagnose tuberculosis (TB) in people with HIV (PWH). We evaluated urine LAM testing conducted in parallel with tests for *Mycobacterium tuberculosis* DNA in oral swabs. In a cohort of 131 South Africans (92% with HIV), combined urine LAM and oral swab testing was significantly more sensitive than either sample tested alone (57% vs. 35% and 39%, respectively), and 97% specific, compared to reference sputum testing (TB culture and Xpert Ultra). Complementary non-sputum sample testing increased sensitivity of TB diagnosis, without sacrificing specificity.

## Background

Tuberculosis disease (TB), caused by *Mycobacterium tuberculosis* (MTB), remains a major cause of illness and death in PWH [1]. The standard sample for TB diagnosis is sputum, which can be difficult for PWH to produce and insensitive in some patients. The availability of noninvasive alternatives to sputum testing would substantially improve TB care for PWH [2, 3].

Immunoassays for mycobacterial lipoarabinomannan (LAM) in urine are potential alternatives to sputum testing. However, the single WHO-approved urine LAM test (by Alere/Abbott) lacks sensitivity. Although next-generation urine LAM tests are in development or evaluation, even best-performing tests of the LAM analyte alone may not be optimally sensitive to detect TB disease in PWH [4, 5].

In search of an alternative non-sputum sample type, we and others have shown that MTB DNA can be detected by oral swab analysis (OSA) [6-14]. In OSA, the dorsum of the tongue is brushed with a sterile swab, and the collected material is tested for MTB DNA. This non-invasive, non-sputum sample collection approach can be applied to any patient, in any setting, including tertiary hospital, outpatient clinic, remote health point, or home and community settings.

Despite their promise, to date neither OSA nor urine LAM testing has proven to be sensitive enough to detect all TB in PWH. We hypothesize that sensitivity is limited for different reasons in the two methods. For example, pulmonary disease may be required to deposit enough MTB in the mouth for detection by OSA, whereas a very high total mycobacterial burden, including extrapulmonary disease, may be required for transrenal passage of MTB glycolipids into urine. Therefore, a parallel approach using both methods may detect more TB cases in PWH than either method alone. The current study was conducted to assess the complementarity of the two methods.

## Methods

### Participant enrollment

South African adults (age ≥16 years) with HIV (regardless of symptoms), or adults with TB symptoms or a positive sputum Xpert Ultra TB test, were consecutively enrolled into the prospective PROVE-TB cohort at Edendale Hospital and affiliated clinics in Pietermaritzburg, South Africa between October 2019 and February 2021. Persons who had received TB trea™ent for more than 24 hours were excluded. Clinical, laboratory, and demographic data were collected from participants and clinical charts. Sputum, urine, tongue swab, and blood samples were collected at bedside and transported to the on-site laboratory for processing and analysis. Study data were collected and managed using REDCap electronic data capture tools hosted at the Institute of Translational Health Sciences. All participants provided written informed consent. This study was approved by the ethical committees of the University of KwaZulu-Natal (BREC #BE475/18) and the University of Washington.

### Sample collection and analysis

Expectorated sputum samples were tested with Cepheid GeneXpert MTB/RIF Ultra (Xpert Ultra) and mycobacterial culture (National Health Laboratory System, South Africa). A TB case was defined as a participant with either a positive sputum Xpert Ultra or a positive sputum TB culture result (reference testing). Spontaneously passed urine samples were tested for the presence of LAM using the Alere DETERMINE LAM Ag lateral-flow assay. A result of 1+ or higher according to the manufacturer’s visual read-out card was considered positive. Two tongue dorsum swabs (Copan FLOQSwabs) were collected into separate tubes as described previously [7, 8]. Swabs in 0.5 mL TE buffer were stored frozen at − 80 °C and transported to the Cangelosi Lab for blinded analysis. Thawed samples were manually processed (Qiagen), concentrated by ethanol precipitation, and tested by IS6110-targeted qPCR. Swab analysis methods were as described previously [7, 8] with the following modifications. Post-boil samples were eluted from the swab head then split, 250 µL reserve and 250 µL for immediate processing by volume-scaled-lysis (300 µL each of Qiagen Buffer AL and 100% ethanol). The qPCR protocol used New England BioLabs® Inc Luna® Universal Probe qPCR Master Mix, and the ethanol-precipitated samples were rehydrated in 5 µL TE buffer prior to master mix addition. Cq values were recorded and results were calculated using two Cq thresholds (38 as described previously [7, 8], and a more stringent value of 32).

In addition, a subset of reserved or duplicate oral swab samples (N=18) were tested using by either of two methods. In one method, previously boiled, reserved (200 µL) half samples were thawed and received 2.2 mL TE buffer. The samples were shaken on a lab vortexer (GENIE® SI-0236 Vortex-Genie 2 Mixer, 120V) on setting 10 for 10-15 seconds, then allowed to sit at ambient temperature for 5 minutes and then shaken for an additional 10-15 seconds, before allowing them sit for additional 10 minutes at ambient temperature. The entire recoverable sample volume was transferred into the sample reservoir of the Xpert Ultra cartridge for analysis. Samples were recorded as positive for MTB if GeneXpert software returned any positive result. Some samples were run by an alternative method that did not use heat. These 0.5-mL samples were thawed at ambient temperature for 30 min, then supplemented with 0.3 mL TE and 1.6 mL of Cepheid Sample Reagent (SR). The samples were vortexed for 10-15 seconds, and incubated for 5 minutes before vortexing for an additional 10-15 seconds. The samples were further incubated for additional 10 minutes before the entire sample volume was transferred to the cartridges.

Sensitivities and specificities of index methods (swabs, urine LAM, and parallel testing) were calculated relative to the reference method (positive sputum Xpert Ultra or positive sputum culture).

## Results

We enrolled 131 participants (45% female, median age 36 years) who provided all sample types for index and reference testing. 120 (92%) participants had HIV. 64 (49%) were TB cases by reference testing **(Figure 1)**.

**Figure 1.**
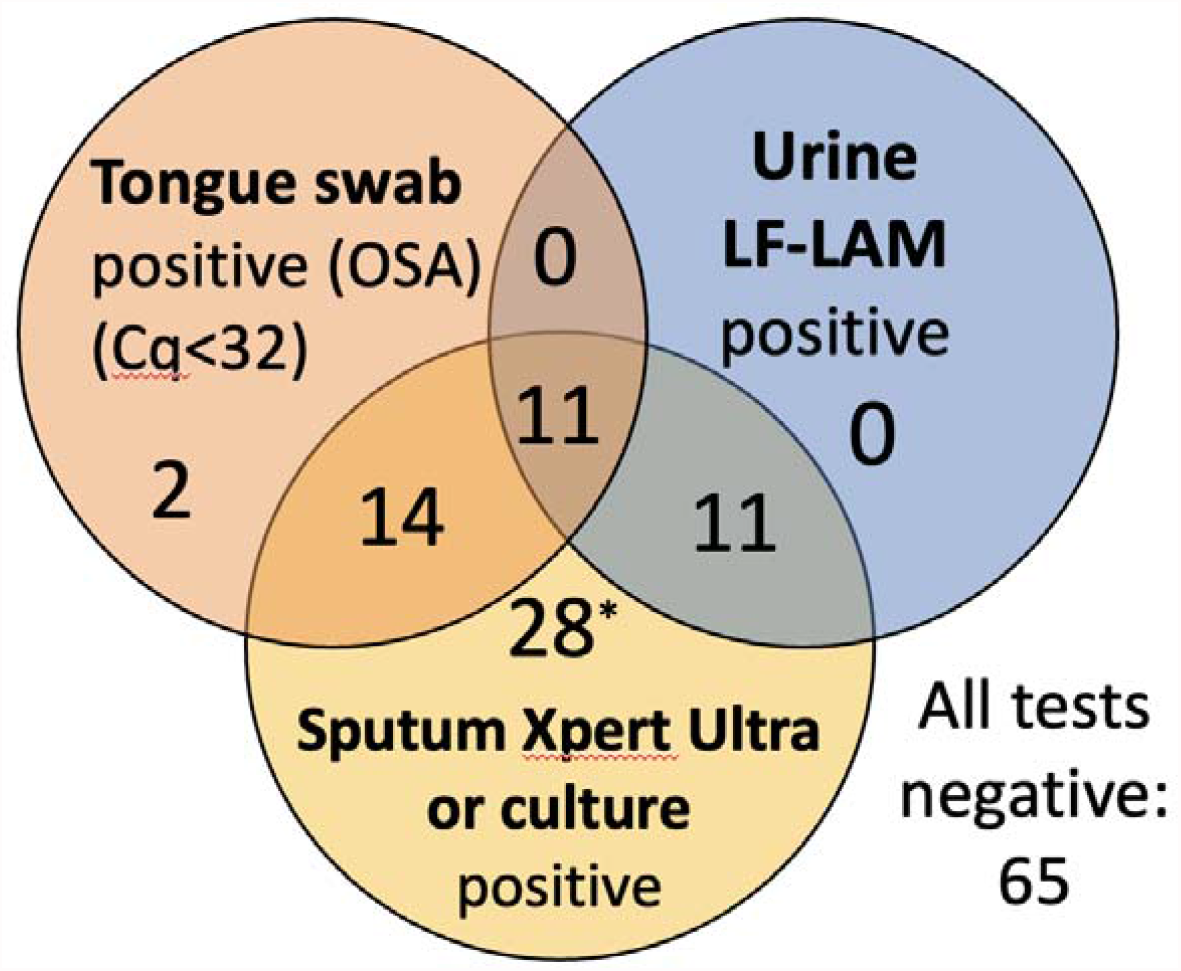
Venn diagram showing overlap of the 2 non-sputum tests (tongue swab and urine LAM) and the sputum TB reference (Xpert Ultra or TB culture). The number of positive participants falling within each category is shown (N=131 participants). *One TB ref+/OSA-participant had an invalid LAM result.

Relative to the sputum reference standard, OSA using manual qPCR conducted on a single swab, with a Cq cutoff of 38, was significantly more sensitive than urine LAM (respectively, 42/64 [67%] vs. 22/63 [35%]). However, OSA was less specific than urine LAM (respectively, 52/67 [78%] vs. 67/67 [100%]). When a more stringent Cq cutoff of 32 was applied to define a positive OSA result, OSA and urine LAM performed similarly (respectively, 25/64 [39%] vs. 22/63 [35%] sensitive, and 65/67 [97%] vs. 67/67 [100%] specific) **(Table 1)**.

**Table 1.** Sensitivity and specificity of non-sputum tests alone or in combination, compared to TB reference standard (sputum Xpert Ultra or sputum culture).

|  | Sensitivity | Specificity |
| --- | --- | --- |
| Tongue Swab (OSA) Cq<32 | 25/64 (39%) | 65/67 (97%) |
| Tongue Swab (OSA) Cq<38 | 42/64 (67%) | 52/67 (78%) |
| Urine LF-LAM | 22/63 (35%) | 67/67 (100%) |
| LAM or OSA*<sub>Cq<32</sub> | 36/63 (57%) | 65/67 (97%) |
| LAM or OSA*<sub>Cq<38</sub> | 45/63 (71%) | 52/67 (78%) |

\* OSA by manual qPCR using 2 thresholds of positivity: Cq<38 (liberal) and Cq<32 (stringent).

When evaluating parallel (combination) non-sputum testing, a positive index test was defined as being <u>either</u> urine LAM-positive <u>or</u> OSA-positive. With parallel testing, sensitivity improved to 36/63 [57%], significantly better than urine LAM alone (p=0.006) or OSA alone (p=0.021). Of the 36 participants who were true positive by non-sputum methods, 11 were positive by both methods while 25 were positive by only one or the other (14 by OSA and 11 by urine LAM; **Figure 1**). Specificity of the combined method remained high at 65/67 (97%).

The manual qPCR method used in Table 1 performed well in past OSA studies [7, 8, 14] but is impractical for routine diagnostic use. Therefore, as a secondary analysis we used the semi-automated Cepheid GeneXpert MTB/RIF Ultra platform to test a subset of 18 frozen swab samples from the current study (11 true positives with strong qPCR signals by OSA, 5 true negatives by OSA, and the 2 false positives by OSA). Xpert Ultra detected 10 of 11 true positives by OSA (the 11^th^ sample had an invalid Xpert Ultra result). It correctly excluded all 5 true negatives by OSA. It also correctly excluded the 2 false positives by OSA **(Table S1)**.

## Discussion

Many PWH struggle to produce sputum (sputum-scarce) and/or have low MTB cell counts in sputum (paucibacillary) [2, 3]. This study tested the hypothesis that parallel testing of two different non-sputum samples can deliver complementary, not redundant diagnostic information, and thereby serve as an alternative to sputum testing for at least some patients.

Recent years have seen advancements in urine LAM testing for sputum-scarce or paucibacillary patients. LAM is an MTB cell wall glycolipid that can pass transrenally into urine. In some patients, most notably PWH with low CD4 counts, LAM is found in the urine in sufficient quantities to enable rapid detection by using lateral flow tests such as the Alere Determine™ TB LAM Ag. However, the sensitivity of urine LAM tests remains suboptimal [4, 5].

We hypothesized that OSA can improve TB diagnosis in paucibacillary and sputum-scarce patients when used in combination with urine LAM testing. The physiological bases for urine LAM testing (transrenal passage of a mycobacterial glycolipid) and OSA (deposition of MTB cells and/or DNA on the tongue dorsum during cough, exhalation, or spontaneous sputum production) are likely to differ from each other. Urine LAM may work best in cases that are at least partially extrapulmonary, while OSA may be best in pulmonary TB. The strengths and limitations of the two methods may therefore complement each other.

The results were consistent with this hypothesis. When the more stringent Cq cutoff of 32 was applied, a diagnostic criterion of positivity in either urine LAM or OSA was significantly more sensitive than either method alone. Specificity was acceptable at 97% and was the minimum of the individual test specificities, but not lower in combination.

This study focused on the biological question of complementarity. Limitations associated with implementation were only partially addressed. While swab sample collection has minimal resource requirements, the manual qPCR method for OSA requires laboratory facilities, specialized equipment and skilled technical operators, so is not practical for routine clinical laboratory use in its current form. However, analysis of a subset of 18 samples with the near point-of-care Xpert Ultra platform suggested that similar results can be obtained with automated tests (with the caveat that the true-positive samples within the subset were strongly positive by manual qPCR). Therefore, this strategy can in theory be applied in any setting with access to both Alere Determine™ TB LAM AG and Xpert Ultra. Other automated amplification platforms intended for near-point-of-care or point-of-care use are also in development and have the potential to be configured for TB OSA.

Another implementation concern is the requirement for two laboratory tests per patient. In context, the same limitation often applies to the current sputum-dependent strategies (paucibacillary and sputum-scarce patients often require multiple sampling and testing attempts to confirm TB). Urine and tongue swabs are easier and safer to collect than sputum samples in most settings. To conserve resources, the two specimens could be collected simultaneously and then tested in a staged reflex algorithm. By design, oral swabs are easy and inexpensive to collect from any patient, in any setting. Swabs can therefore be collected and stored concurrently with the collection of all urine samples. If a patient’s urine sample is positive for LAM, then that is diagnostic for TB and a swab test is not needed. If the urine LAM test is negative, then the swab can be tested to avoid having to recall the patient for sputum induction.

In conclusion, our results indicate that tongue swabs and urine can serve as complementary non-sputum samples for improved diagnosis of TB in PWH. With further optimization to improve sensitivity, this approach may be considered as a fully non-sputum strategy for detecting TB in sputum-scarce patients.

## Data Availability

All data produced in the present study are available upon reasonable request to the authors

## Footnote page

### Conflict of interest statement

The authors have no conflicts of interest to declare.

This work was supported by the Bill and Melinda Gates Foundation (#INV-004527, OPP 1213054), by NIH grants R01AI139254 and U54EB027049. REDCap at ITHS is supported by the National Center For Advancing Translational Sciences of the National Institutes of Health under Award Number UL1 TR002319. AES is supported by NIH (NIAID) K23A140918. We are grateful to Cepheid, Inc., for providing research-use-only Xpert Ultra test kits, and to Copan Italia for providing swabs.

This work was presented in part at virtual TB Science 2021, (Abstract #TBS-LB-2021-02134)

## Literature cited

1. World Health Organization. Global Tuberculosis Report 2020.

2. UNITAID. Tuberculosis diagnostics technology and market landscape - 5th edition. World Health Organization. 2017.

3. Fauci, AS, Eisinger RW. Reimagining the Research Approach to Tuberculosis. Am J Trop Med Hyg. 2018;98(3):650–2.

4. Shah M, Hanrahan C, Wang ZY, Dendukuri N, Lawn SD, Denkinger CM, et al. Lateral flow urine lipoarabinomannan assay for detecting active tuberculosis in HIV-positive adults. Cochrane Database of Systematic Reviews. 2016;(5).

5. Broger T, Sossen B, du Toit E, Kerkhoff AD, Schutz C, Ivanova Reipold E, et al. Novel lipoarabinomannan point-of-care tuberculosis test for people with HIV: a diagnostic accuracy study. The Lancet Infectious Diseases. 2019;19(8):852–61.

6. LaCourse SM, Seko E, Wood R, Bundi W, Ouma GS, Agaya J, et al. Diagnostic performance of oral swabs for non-sputum based TB diagnosis in a TB/HIV endemic setting. PLoS One. 2022;17(1):e0262123. Epub 2022/01/14.

7. Wood RC, Andama A, Hermansky G, Burkot S, Asege L, Job M, et al. Characterization of oral swab samples for diagnosis of pulmonary tuberculosis. PLOS ONE. 2021;16(5):e0251422. doi: 10.1371/journal.pone.0251422.

8. Luabeya AK, Wood RC, Shenje J, Filander E, Ontong C, Mabwe S, et al. Noninvasive Detection of Tuberculosis by Oral Swab Analysis. Journal of Clinical Microbiology. 2019;57(3):e01847–18.

9. Wood RC, Luabeya AK, Weigel KM, Wilbur AK, Jones-Engel L, Hatherill M, et al. Detection of Mycobacterium tuberculosis DNA on the oral mucosa of tuberculosis patients. Scientific Reports. 2015;5:8668.

10. Nicol MP, Wood RC, Workman L, Prins M, Whitman C, Ghebrekristos Y, et al. Microbiological diagnosis of pulmonary tuberculosis in children by oral swab polymerase chain reaction. Scientific Reports. 2019;9(1):10789.

11. Valinetz ED, Cangelosi GA. A Look Inside: Oral Sampling for Detection of Non-Oral Infectious Diseases. Journal of Clinical Microbiology. 2021:JCM.02360-20.

12. Flores JA, Calderon R, Mesman AW, Soto M, Coit J, Aliaga J, et al. Detection of Mycobacterium Tuberculosis DNA in Buccal Swab Samples from Children in Lima, Peru. The Pediatric Infectious Disease Journal. 2020;39(11).

13. Mesman AW, Calderon R, Soto M, Coit J, Aliaga J, Mendoza M, et al. Mycobacterium tuberculosis detection from oral swabs with Xpert MTB/RIF ULTRA: a pilot study. BMC Res Notes. 2019;12(1):349-.

14. Song Y, Ma Y, Liu R, Shang Y, Ma L, Huo F, et al. Diagnostic Yield of Oral Swab Testing by TB-LAMP for Diagnosis of Pulmonary Tuberculosis. Infect Drug Resist. 2021;14:89–95.

