## Supplementary material for "Complementary non-sputum diagnostic testing using oral swabs and urine LAM testing for TB in people with HIV": Table S1

**Supplementary data**

**Table S1. Diagnostic performance of Xpert Ultra and manual qPCR on 18 tongue swab samples, compared to TB reference sputum testing (ref).**

| **Tongue swab subset (N=18)** | **TB ref* pos** | **TB ref neg** |
| --- | --- | --- |
| OSA Xpert Ultra pos  OSA Xpert Ultra neg | 10**  0 | 0  7 |
| OSA qPCR_Cq<32_ pos  OSA qPCR_Cq<32_ neg | 11  0 | 2  5 |

*Ref: sputum Xpert Ultra or culture positive. Pos: positive. Neg: negative

**One patient with TB ref+ sputum sample had an invalid result by OSA Xpert Ultra.
